# Racial identity and mental health stigma among Black adults in the United States

**DOI:** 10.1101/2021.10.27.21265589

**Authors:** Aderonke Bamgbose Pederson, Devan Hawkins

## Abstract

**Objectives:** Mental illness stigma is a barrier to engagement in mental health services. This study assesses our hypothesis that specific racial identity dimensions influences mental health behavior including stigma.

**Methods:** We performed an online cross sectional observational study among Black adults (n = 248, ages 18-65). We examined the relationship between an individual’s approach to their racial identity in the community and stigma behavior towards mental health; generalized linear models were performed. We assessed demographic characteristics as moderators of the primary association.

**Results:** Black adults with higher centrality reported lower past stigma behavior (RR=1.57, CI: 1.11–2.21, p=0.01), but higher future intended stigma behavior (RR=0.93, CI: 0.88–0.99, p=0.02). Majority of respondents reported high centrality and high assimilation; however, assimilation did not appear to correlate with mental health stigma behavior. Age, education and ethnicity appeared to have a limited moderating effect on the association between centrality and stigma behavior.

**Conclusions:** Centrality was associated with mental health stigma behavior. By understanding the intersecting characteristics that may increase the likelihood for mental illness stigma, we will be better able to reduce mental illness stigma and optimize engagement in mental health services.

## Introduction

Black people continue to experience mental health disparities in the United States (1, 2). Disparities among Black people result in low engagement in mental health services, more chronic disease, higher levels of disability, higher rates of inpatient hospitalizations and lower rates of outpatient mental health service use compared to their White people (1, 3-5). Several factors contribute to health disparities including stigma related to mental illness, which is a leading cause of health inequities (6). Black people hold stigmatizing views towards mental illness and utilization of mental health services (1, 3-5). Mental illness stigma results in barriers to mental health service delivery among Black people. Stigma refers to the negative attitudes and beliefs one may hold towards mental illness and is based on a desire for social distance or separation from people who are part of the stigmatized group (7-9).

We focus on racial identity among African Americans and Black immigrants and how one’s interaction with their racial identity and community influences mental health behavior (10). Some studies show greater connection to the mainstream culture improves mental health while other studies show greater connection to mainstream culture (or lower connection to one’s racial community) contributes to greater psychological distress (11, 12). Therefore, it is important to understand how racial identity in the Black community may influence mental health behaviors such as stigma related behaviors, given its known influence on mental health outcomes. This association is understudied among Black adults including Black immigrants and African Americans.

We focus on centrality and assimilation aspects of Black identity in the U.S. We assessed social distance from people with mental illness, which is an indicator of past, current or future discriminatory manifestations of mental illness stigma (13, 14). We assessed centrality, which refers to the extent to which being Black is core to one’s self concept; we also assessed assimilation, which refers to the extent to which Black people should strive to be integrated into the mainstream culture (15). In this study, we examined the relationship between racial identity (using centrality and assimilation factors) and social distance from persons with mental health problems. We adjusted for ethnicity between those who identified as African Americans and those who identified as Black immigrants, given the salience of migration and Black identity in the U.S.

## Methods

### Study design

We performed a cross-sectional observational study to assess characteristics of racial identity and stigma among Black adults who identify as African-American, African immigrants and Afro-Caribbean immigrants from September 2020 to October 2020.

### Participants and study setting

The study participants were recruited from community based organizations serving Black adults and represent a convenience sample (n = 248). The online survey was shared using flyers and social media platforms. Eligibility criteria included ages 18-65, identifying as Black, African-American, African or Afro-Caribbean; currently living in the U.S.; and English speaking.

### Procedure

The self-report surveys were distributed using Qualtrics software. This study reports on the data collected regarding the association between racial identity and mental illness stigma behavior.

### Ethical Review

The Northwestern University Institutional Review Board approved this study prior to data collection.

### Measures

We collected data using validated and standardized instruments to measure Black racial identity salience. We also measured stigma related to mental health. Participants self-report on how they interact with their racial identity was based on two subscales in the Multidimensional Inventory of Black Identity (MIBI). The subscales included measures on centrality and assimilation. We also included a measure of Reported and Intended Behavior Scale (RIBS) to assess current or past and future behavior related to people with mental health problems. The demographic questionnaire: The demographic questionnaire included self-report questions on age, education, gender, ethnicity, marital status, income, and insurance status.

The MIBI (Multidimensional Inventory of Black Identity): a 56-item racial and ethnic identity survey (16): The MIBI has been validated among Black people across age groups living in the U.S. as well as among Black people in other countries. We focused on two subscales within this measure, an 8-item centrality subscale and a 9-item assimilation subscale. The centrality dimension assesses the extent to which individuals believe being Black is central to their self-definition or identity, for example, “in general, being Black is an important part of my self-image.” The assimilation subscale assesses the extent to which Black people should strive to be integrated into the mainstream culture and political system and focuses on the commonalities between Black people and other Americans. The scales have moderate to high internal consistency and the cronbach’s alphas for each subscale ranged from 0.60 to 0.80 (16, 17). The RIBS (Reported and Intended Behavior Scale) (14): The RIBS is an 8-item scale divided into two sections (reported past and current behavior related to mental health problems and reported future intended behavior). There are 4-items in each section. Each of the first four items are examined as individual outcomes with responses yes or no or don’t know, representing the prevalence of a particular behavior in the past or current time. The second four items are assessed as a total score (4-20) based on responses on an ordinal scale (1-5), a higher score indicates greater willingness to engage in a particular behavior and hence less future intended stigmatizing behavior. Each question begins with “in the future I would be willing to⃛” “live,” “work,” “be a neighbor to” or “have a close friendship with someone with a mental health problem”. The scale showed good validity (cronbach’s alpha 0.72 to 0.81 for the 4-item individual measures and 0.85 for the subscale on future intended behavior) and good reliability (test-retest reliability was 0.75) (14).

### Data analysis

The frequency and percentage of participants according to age group, gender, education, income, ethnicity, and citizenship status were calculated. In order to examine the relationship between the approach to Black identity within the U.S. (MIBI) and stigma behavior towards mental health (RIBS), generalized linear models in SAS Version (9.3) were used. For the relationship between the centrality and assimilation dimensions of Black identity and the first four items of the RIBS scale, Poisson regression with a log link was used to calculate rate ratios (RR) in order to assess the changes in the probability of responding ‘yes’ to the RIBS items per unit change in mean score for each of the racial identity dimensions. When the total RIBS score (future intended stigmatizing behavior based on the second four items) was treated as an outcome, gamma regression using generalized linear models in SAS Version (9.3) was used. The RRs from these models represent the average change in the RIBS score per unit change in the centrality and assimilation dimensions.

To examine whether these findings were potentially confounded, models were constructed that controlled for age, education and ethnicity. Finally, to explore whether there was effect modification according to these variables, these same regression models were run while stratified according to age, education and ethnicity. Afro-Caribbean immigrants (n=11) were combined with African immigrants (n=47) into a *Black immigrant* group due to their small sample size. There is precedent for this, given some shared characteristics of migration experienced by immigrants and its intersection with race i.e. identifying as Black (18).

## Results

As shown in table 1, among the 248 participants, 59.27 % of participants were aged 18-34. Some participants (30.24%) reported having no college education. Some participants reported a personal income less than $30, 000 (43.15%). Over half of the participants reported having public health insurance (55.23%).

**Table 1.** Descriptive demographics and general percentages for the sample

|  | n | % |
| --- | --- | --- |
| Age |  |  |
| 18-34 | 147 | 59.27 |
| 35-65 | 101 | 40.73 |
| Gender |  |  |
| Male | 141 | 56.85 |
| Female | 107 | 43.15 |
| Marital status |  |  |
| Married | 158 | 63.71 |
| Unmarried, living with a romantic | 41 | 16.53 |
| partner |  |  |
| Never married | 26 | 10.48 |
| Separated | 7 | 2.82 |
| Divorced | 15 | 6.05 |
| Widowed | 1 | 0.40 |
| Education |  |  |
| Less than high school | 4 | 1.61 |
| High school diploma / GED | 38 | 15.32 |
| Trade school/vocational school | 33 | 13.31 |
| Some college, no degree | 85 | 34.27 |
| 2 year college | 42 | 16.94 |
| 4-year college degree | 40 | 16.13 |
| Master's degree | 4 | 1.61 |
| Doctoral degree | 2 | 0.81 |
| Income |  |  |
| \$9,999 or less | 14 | 5.65 |
| \$10,000 to \$29,999 | 93 | 37.50 |
| \$30,000 to \$49,999 | 108 | 43.55 |
| \$50,000 to \$69,999 | 17 | 6.85 |
| \$70,000 to \$89,999 | 7 | 2.82 |
| \$90,000 to \$109,999 | 4 | 1.61 |
| \$110,000 to \$129,999 | 3 | 1.21 |
| .\$130, 000 to \$189,999 | 0 | 0.00 |
| \$190,000 to \$199,999 | 1 | 0.40 |
| \$200,000 or more | 1 | 0.40 |
| Health Insurance |  |  |
| None | 59 | 23.79 |
| Public Insurance | 132 | 53.23 |
| Private | 57 | 22.98 |
| Ethnicity |  |  |
| African | 47 | 18.95 |
| African American | 190 | 76.61 |
| Afro-Caribbean | 11 | 4.44 |
| Citizenship status |  |  |
| Citizen | 188 | 75.81 |
| Non-citizen | 60 | 24.19 |

### Characteristics of participants as measured by the MIBI

In response to questions on racial identity, three-fifth (59.7%) of respondents endorsed that being Black was an important part of their self-image and 60.3% of respondents endorsed Black people should strive to integrate all institutions, which are segregated. We report on several other racial identity characteristics of the sample in table 2.

**Table 2.** Racial identity characteristics among participants

| **Racial identity characteristics** | **Percent of sample** |
| --- | --- |
| **Sample centrality responses** |  |
| • Respondents endorsed that being Black was an important part of their self-image | 59.7% |
| • Respondents endorsed that their destiny was tied to the destiny of other Black people | 57.0% |
| • Respondents endorsed that being Black was an important reflection of who they were | 60.7% |
| **Sample assimilation responses** |  |
| • Respondents endorsed Black people should strive to be part of the American political system | 67.0% |
| • Respondents endorsed Black people should work within the system to achieve their economic and political goals | 66.5% |
| • Respondents endorsed Black people should strive to integrate all institutions which are segregated | 60.3% |
| • Respondents endorsed the plight of Black people in America will improve when Black people are in important positions within the system | 62.9% |

### Main analysis and adjusted analysis: centrality and assimilation and its association with reported social distance and future intended stigma behavior

*Association between centrality, assimilation and social distance from people with a mental health problem (see Table 3)*.

**Table 3.** Association between Black identity (centrality and assimilation dimensions) and stigma behavior (reported past, current and intended future behavior)

|  | Reported social distance: living with PWMI |  |  |  | Reported social distance: working with PWMI |  |  |  | Reported social distance: neighbor with PWMI |  |  |  | Reported social distance: close friend with PWMI |  |  |  | Future intended stigma behavior (RIBS average) |  |  |  |
| --- | --- | --- | --- | --- | --- | --- | --- | --- | --- | --- | --- | --- | --- | --- | --- | --- | --- | --- | --- | --- |
|  | Model 1 |  | Model 2 |  | Model 1 |  | Model 2 |  | Model 1 |  | Model 2 |  | Model 1 |  | Model 2 |  | Model 1 |  | Model 2 |  |
|  | RR (95% CI) | P-value | RR (95% CI) | P-value | RR (95% CI) | P-value | RR (95% CI) | P-value | RR (95% CI) | P-value | RR (95% CI) | P-value | RR (95% CI) | P-value | RR (95% CI) | P-value | RR (95% CI) | P-value | RR (95% CI) | P-value |
| Centrality | **1.39 (1.06, 1.82)** | **0.0189\*** | **1.37 (1.03, 1.81)** | **0.0285\*** | 1.05 (0.81, 1.37) | 0.72 | 1.12 (0.85, 1.49) | 0.4151 | 1.20 (0.90, 1.60) | 0.2095 | 1.17 (0.88, 1.57) | 0.2717 | 1.20 (0.89, 1.60) | 0.2291 | 1.23 (0.91, 1.66) | 0.1787 | 0.96 (0.91, 1.01) | 0.0951 | 0.96 (0.91, 1.01) | 0.1040 |
| Assimilation | 1.21 (0.95, 1.56) | 0.1223 | 1.20 (0.94, 1.54) | 0.1491 | 1.04 (0.83, 1.30) | 0.7577 | 1.07 (0.84, 1.36) | 0.5664 | 1.24 (0.96, 1.60) | 0.106 | 1.24 (0.95, 1.60) | 0.1091 | 1.08 (0.84, 1.40) | 0.5338 | 1.09 (0.84, 1.41) | 0.5309 | 0.98 (0.94, 1.02) | 0.3089 | 0.98 (0.94, 1.02) | 0.3921 |

Poisson regression with a log link was used to calculate rate ratios (RR) in order to assess the changes in the probability of social distance and stigma per unit change in mean score for each of the centrality and assimilation components of Black identity. For future intended stigmatizing behavior, gamma regression using generalized linear models in SAS Version (9.3) was used. These models represent the average change in the RIBS score per unit change in the centrality and assimilation dimensions. Model 1 – Only acculturation; Model 2 – adjustment for age, education, and ethnicity. PWMI: person living with a mental illness. **\*statistically significant outcomes $p < 0.05$ .**

Respondents who reported higher centrality were more likely to report living with or having ever lived with someone with a mental health problem. The probability of reporting living with or having ever lived with someone with a mental health problem increased by 39% per unit increase in the centrality score (RR = 1.39, CI: 1.06 – 1.82, p = 0.019). After adjusting for age, education and ethnicity, there was a 37% change per unit increase in the centrality score (RR = 1.37, CI: 1.03 – 1.82, p = 0.028).

*Association between centrality, assimilation and social distance from people with a mental health problem, findings on effect modification of age, education and ethnicity (see Table 4)*

**Table 4.**
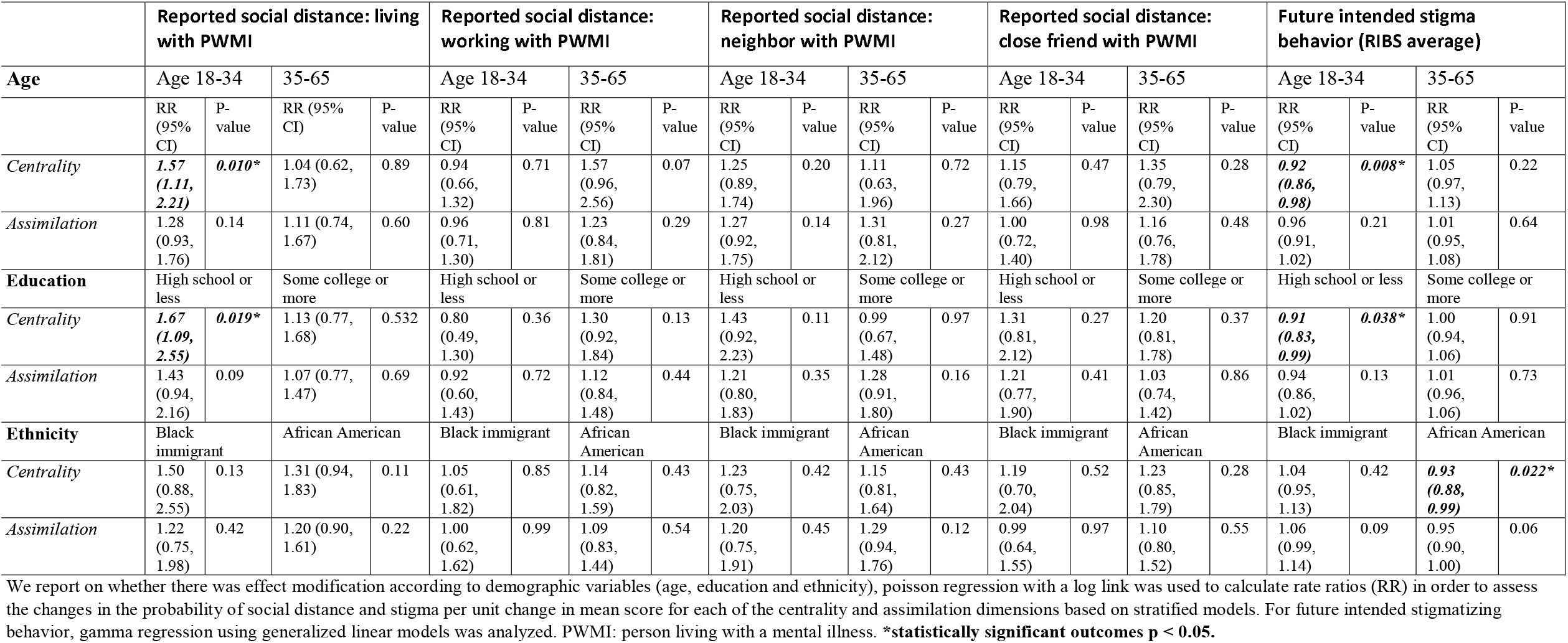
Association between Black identity (centrality and assimilation dimensions) and stigma behavior (reported past, current and intended future behavior) with effect modification by age, education, and ethnicity

|  | Reported social distance: living with PWMI |  |  |  | Reported social distance: working with PWMI |  |  |  | Reported social distance: neighbor with PWMI |  |  |  | Reported social distance: close friend with PWMI |  |  |  | Future intended stigma behavior (RIBS average) |  |  |  |
| --- | --- | --- | --- | --- | --- | --- | --- | --- | --- | --- | --- | --- | --- | --- | --- | --- | --- | --- | --- | --- |
| Age | Age 18-34 |  | 35-65 |  | Age 18-34 |  | 35-65 |  | Age 18-34 |  | 35-65 |  | Age 18-34 |  | 35-65 |  | Age 18-34 |  | 35-65 |  |
|  | RR (95% CI) | P-value | RR (95% CI) | P-value | RR (95% CI) | P-value | RR (95% CI) | P-value | RR (95% CI) | P-value | RR (95% CI) | P-value | RR (95% CI) | P-value | RR (95% CI) | P-value | RR (95% CI) | P-value | RR (95% CI) | P-value |
| *Centrality* | **1.57 (1.11, 2.21)** | **0.010\*** | 1.04 (0.62, 1.73) | 0.89 | 0.94 (0.66, 1.32) | 0.71 | 1.57 (0.96, 2.56) | 0.07 | 1.25 (0.89, 1.74) | 0.20 | 1.11 (0.63, 1.96) | 0.72 | 1.15 (0.79, 1.66) | 0.47 | 1.35 (0.79, 2.30) | 0.28 | **0.92 (0.86, 0.98)** | **0.008\*** | 1.05 (0.97, 1.13) | 0.22 |
| *Assimilation* | 1.28 (0.93, 1.76) | 0.14 | 1.11 (0.74, 1.67) | 0.60 | 0.96 (0.71, 1.30) | 0.81 | 1.23 (0.84, 1.81) | 0.29 | 1.27 (0.92, 1.75) | 0.14 | 1.31 (0.81, 2.12) | 0.27 | 1.00 (0.72, 1.40) | 0.98 | 1.16 (0.76, 1.78) | 0.48 | 0.96 (0.91, 1.02) | 0.21 | 1.01 (0.95, 1.08) | 0.64 |
| **Education** | High school or less |  | Some college or more |  | High school or less |  | Some college or more |  | High school or less |  | Some college or more |  | High school or less |  | Some college or more |  | High school or less |  | Some college or more |  |
| *Centrality* | **1.67 (1.09, 2.55)** | **0.019\*** | 1.13 (0.77, 1.68) | 0.532 | 0.80 (0.49, 1.30) | 0.36 | 1.30 (0.92, 1.84) | 0.13 | 1.43 (0.92, 2.23) | 0.11 | 0.99 (0.67, 1.48) | 0.97 | 1.31 (0.81, 2.12) | 0.27 | 1.20 (0.81, 1.78) | 0.37 | **0.91 (0.83, 0.99)** | **0.038\*** | 1.00 (0.94, 1.06) | 0.91 |
| *Assimilation* | 1.43 (0.94, 2.16) | 0.09 | 1.07 (0.77, 1.47) | 0.69 | 0.92 (0.60, 1.43) | 0.72 | 1.12 (0.84, 1.48) | 0.44 | 1.21 (0.80, 1.83) | 0.35 | 1.28 (0.91, 1.80) | 0.16 | 1.21 (0.77, 1.90) | 0.41 | 1.03 (0.74, 1.42) | 0.86 | 0.94 (0.86, 1.02) | 0.13 | 1.01 (0.96, 1.06) | 0.73 |
| **Ethnicity** | Black immigrant |  | African American |  | Black immigrant |  | African American |  | Black immigrant |  | African American |  | Black immigrant |  | African American |  | Black immigrant |  | African American |  |
| *Centrality* | 1.50 (0.88, 2.55) | 0.13 | 1.31 (0.94, 1.83) | 0.11 | 1.05 (0.61, 1.82) | 0.85 | 1.14 (0.82, 1.59) | 0.43 | 1.23 (0.75, 2.03) | 0.42 | 1.15 (0.81, 1.64) | 0.43 | 1.19 (0.70, 2.04) | 0.52 | 1.23 (0.85, 1.79) | 0.28 | 1.04 (0.95, 1.13) | 0.42 | **0.93 (0.88, 0.99)** | **0.022\*** |
| *Assimilation* | 1.22 (0.75, 1.98) | 0.42 | 1.20 (0.90, 1.61) | 0.22 | 1.00 (0.62, 1.62) | 0.99 | 1.09 (0.83, 1.44) | 0.54 | 1.20 (0.75, 1.91) | 0.45 | 1.29 (0.94, 1.76) | 0.12 | 0.99 (0.64, 1.55) | 0.97 | 1.10 (0.80, 1.52) | 0.55 | 1.06 (0.99, 1.14) | 0.09 | 0.95 (0.90, 1.00) | 0.06 |

We report on whether there was effect modification according to demographic variables (age, education and ethnicity), poisson regression with a log link was used to calculate rate ratios (RR) in order to assess the changes in the probability of social distance and stigma per unit change in mean score for each of the centrality and assimilation dimensions based on stratified models. For future intended stigmatizing behavior, gamma regression using generalized linear models was analyzed. PWMI: person living with a mental illness. **\*statistically significant outcomes p < 0.05.**

We assessed age, education and ethnicity as moderators of the relationship between Black identity dimensions (centrality and assimilation) and social distance from persons with mental illness in the past, current and future time. We found statistically significant association between centrality and social distance and stigma related behavior. However, in our analysis of effect modification, we did not find a statistically significant correlation between assimilation and social distance or stigmatizing behavior.

In our assessment of age as a moderator, we found that for respondents who are less than 35 years old, the probability of reporting living with or having ever lived with someone with a mental health problem increased by 57% (RR = 1.57, CI: 1.11 – 2.21, p = 0.01) per unit increase in the centrality score. Despite higher likelihood to report living with someone with a mental health problem, this group of respondents who are less than 35 years old, the probability of reporting living with or having ever lived with someone with a mental health problem decreased by 8% (RR = 0.92, CI: 0.86 – 0.98, p = 0.008) per unit increase in the centrality score, hence there was greater future intended stigmatizing behavior as centrality increased.

In our assessment of education as a moderator, we found that for respondents who report having no college education, the probability of reporting living with or having ever lived with someone with a mental health problem decreased by 67% (RR = 1.67, CI: 1.09 – 2.55, p = 0.019) per unit increase in the centrality score. Again despite the likelihood to report living with someone with a mental health problem, respondents who report having no college education, the probability of reporting living with or having ever lived with someone with a mental health problem decreased by 9% (RR = 0.91, CI: 0.83 – 0.99, p = 0.038) per unit increase in the centrality score, hence there was greater future intended stigmatizing behavior as centrality increased.

Finally in our moderation analysis, for respondents who reported identifying as African American, the probability of reporting living with report living with or having ever lived with someone with a mental health problem decreased by 7% (RR = 0.93, CI: 0.88 – 0.99, p = 0.022) per unit increase in the centrality score, hence there was greater future intended stigmatizing behavior as centrality increased. Among African Americans, higher centrality was correlated with greater future intended stigmatizing behavior towards people with mental illness.

## Discussion

We found an association between centrality and social distance from people with a mental health problem; those who endorsed high centrality were less likely to endorse social distance in reference to living with or having ever lived with someone with a mental health problem. Having high centrality (strong identity and self-concept in being a part of the Black community) was associated with social proximity in the past or current time with someone with a mental health problem. Several studies have shown one’s interaction with their racial and ethnic identity (as a component of acculturation) is related to mental health and well-being, however few studies assess how this interaction may be associated with social distance and stigmatizing behaviors from people with a mental illness (19-21). Previous studies have shown high centrality was associated with less depressive symptoms and improved psychological health (17). However, the process by which racial identity is associated with mental health is complex (17, 22). One hypothesis, the insulation hypothesis, proposes that Black people determine their worth and behavior, including mental health behavior, based on comparing themselves to other Black people (15, 17, 23). Our study supports the current literature that indicates there is an association between centrality and mental health behavior, but offers additional specificity to the growing body of knowledge around racial and ethnic identity and mental health behaviors. We found that respondents with higher centrality reported greater social proximity (or less social distance) to people with a mental health problem, even after controlling for demographic variables including ethnicity.

We found an association between centrality and social distance as well as stigmatizing behavior for respondents who were younger (less than 35 years old). Those who were less than 35 years old and reported higher centrality were more likely to report living with someone with mental health problems but they were also more likely to report greater future intended stigmatizing behavior. Hence, past or current experience living with someone with mental health problems did not confer lower future stigmatizing behavior for young Black adults (ages 18 – 34). From our study it appears that high centrality was associated with higher future intended stigma behavior for this age group. Previous studies show that racial and ethnic identity and age are associated in mental health behavior, one study found an association between acculturation and suicidal ideation among young Black adults, they found a 7% increase in suicidal ideation was associated with increase in acculturation (24). Our study contributes additional knowledge around the interaction of racial identity and mental health behavior among younger Black adults, our study suggests that one possible reason greater centrality may lead to negative mental health behavior among young Black adults is because of the positive correlation of centrality and future intended stigmatizing behavior towards mental health.

The relationship seen with younger Black adults was similar to the relationship seen with educational status. Black adults without a college degree who reported high centrality were more likely to report living with someone with mental health problems, however this same group were also more likely to endorse future intended stigmatizing behavior. Previous studies have shown that education reduces mental illness stigma, however one study suggests the opposite, that greater educational attainment may predispose one to greater stigma towards mental illness due to the intersection of race and mental health (25, 26). Greater stigma among those with lower educational attainment with high centrality may be a result of intersecting stigmas such as the stigma that may be associated with lower educational attainment, the stigma that may be associated with having a mental illness and the discrimination associated with being Black (27, 28). Therefore, these interesting stigmas may lead to a desire to minimize additional stigmatization by avoiding future contact with people with mental illness as seen in our study.

Those who identified as African Americans and reported higher centrality were more likely to endorse future intended stigmatizing behavior. Previous studies show that stronger ethnic identity is associated with lower likelihood of using mental health services among Black people (22). A previous study also showed that Black people with higher centrality who reported discriminatory experiences or racism were less likely to use mental health services (22, 29). We found a correlation among African Americans with high centrality and mental health stigma but we did not see a similar relationship among those who identified as Black immigrants, this difference could be related to differing experiences of discrimination and the role of intersectionality specific to immigrant experiences among Black immigrants compared to African Americans (27).

### Study Limitations

Future studies would benefit from a longitudinal analysis to assess causality, we were limited by a cross-sectional design. In cases where multiple comparisons were performed, a bonferroni correction on the p-values may be indicated to assess for chance findings in future studies.

## Conclusion

Our study provides critical data that improves our understanding of the association between racial identity within the Black community and mental health outcomes. Beyond the direct effect of centrality and assimilation on mental health behaviors or outcomes, consideration of mental illness stigma is essential to further describe mental health behavior related to how Black people approach their racial within the U.S. There was a consistent discrepancy between past or current proximity to people with a mental health problem and future intended stigmatizing behavior for people with higher centrality. In addition, age, education and ethnicity were found to influence the relationship between centrality and social distance from people with a mental health problem both in the past, current and future time. We highlight important concepts and lay a foundation towards future studies focused on the mechanistic relationships between centrality, assimilation and stigma. This has implications for the implementation of programs to address mental health disparities.

## Data Availability

All data produced in the present work are contained in the manuscript

## Acknowledgments

The study received funding through the National Center for Advancing Translational Sciences (NCATS), an institute of the National Institute of Health.

The authors have no conflict of interest to disclose

